# Expression patterns of *HMGA2* in the placenta during pregnancy

**DOI:** 10.1101/2020.12.07.20245092

**Authors:** Lars Burchardt, Andrea Gottlieb, Burkhard M. Helmke, Werner Wosniok, Wolfgang Kuepker, Jörn Bullerdiek

## Abstract

**Background:** *High-mobility group AT-hook 2 (HMGA2)* expression can be detected in many embryonic and fetal tissues but becomes down-regulated during postnatal life except for many benign and malignant tumors. In the latter case, its expression has been correlated with epithelial-mesenchymal transition and invasive growth. The placenta contributes essentially to proper development of the embryo and the fetus. In a tumor-like manner it shows rapid invasive growth during the first weeks of gestation. To address the possible role of HMGA2 during placental development, we have measured its expression throughout the prenatal period and in term placentae by mRNA quantification as well as by immunohistochemistry.

**Methods:** Expression of *HMGA2* and *HPRT* was measured on 89 fetal placentas, encompassing calendar gestational age of five to 41 weeks, using quantitative real time-PCR. In eleven cases in addition immunohistochemistry was used to determine the localization of HMGA2 and to compare with data obtained by quantitative real time-PCR.

**Results:** The expression of *HMGA2* was found to be inversely correlated with gestational age (*p* < 0.001). For the first part of the first trimester the level of *HMGA2* is high. After that the expression shows a decline down to a baseline level where it remains until birth. HMGA2 protein was mainly detected in the nuclei of the stromal cells in the placental villi.

**Conclusions:** During pregnancy, the expression of *HMGA2* follows a non-linear pattern of decrease. In the first trimester, from two to three weeks after the implantation of the conceptus until the blood supply is established (hypoxic phase), the expression is high, indicating a critical role during early development and in the control of its invasive behavior, respectively.

## Introduction

Human high-mobility group AT-hook protein 2 belongs to a family of non-histone chromatin proteins encoded by two genes, *HMGA1* and *HMGA2*. Due to alternative splicing, there are four known proteins (HMGA1a, HMGA1b, HMGA1c, and HMGA2) [reviewed in [1]]. All HMGA proteins are architectural transcription factors and contain three DNA-binding domains called AT-hooks and an acidic carboxy-terminal tail. As such, they do not have an intrinsic transcription factor capacity but rather enhance or silence transcription through a change in chromatin structure and interaction with nuclear proteins [reviewed in [2,3]. They play a key role in stem cell renewal, growth and development of tissues, and the differentiation of cells [4-6]. As to the expression of HMGA2 in mammalian cells and tissues, three fields can be distinguished: First, in contrast to embryonic and fetal development its expression is not detectable in most adult tissues and cells [7-10] with few exceptions as e.g. some types of mesenchymal stem cells [11], spermatocytes, and spermatides [12] and constitutional knock-out of *HMGA2* is associated with a pygmy phenotype in mice [13]. Secondly, once activated by chromosomal translocations, it apparently causally can contribute to the growth of a variety of benign tumors of adult tissues such as uterine leiomyomas, lipomas, pulmonary chondroid hamartomas, and pleomorphic adenomas of the salivary glands [14-16]. Thirdly, an overexpression of HMGA2 occurring independent of chromosomal translocations affecting its gene locus and surrounding sequences was detected in numerous types of cancers (reviewed in [1,17,18]) where its expression is usually associated with a higher degree of malignant behavior and worse prognosis, respectively [19-27]. Accordingly, the expression of HMGA2 is considered being a promising universal tumor marker for prognostics [26].

The placenta is the only normal human organ infiltrating surrounding tissue albeit as a rule in a tightly restricted manner following mechanisms that are not fully understood until today. It serves as the connection between mother and embryo or fetus providing nutrients and oxygen for the developing child. About six days after fertilization the blastocyst starts implantation into the maternal uterus and placentation. Five weeks after conception the basic structure of the placenta has formed. To support the developing embryo and fetus it continues to grow throughout gestation. Disturbances of the depth of infiltration are associated with serious problems such as preeclampsia and placenta accrete or percreta.

The detection of *HMGA2* expression in the placenta dates back to 1996 [8]. To detect *HMGA2* mRNA, several embryonal/fetal tissues of a gestational age between eight to twelve weeks and samples of the maternal and the fetal part of a placenta (36th weeks of pregnancy) were analyzed by RT-PCR. The experiments yielded positive results for the embryo but negative findings for the placenta. In an investigation by Hirning-Folz *et al*. [10] RNA in situ hybridization was used on mouse embryo sections for detection of *HMGA2* mRNA. Fetal placenta showed reduced expression compared to nearly all parts of the embryo at a developmental stage of 9.5. *HMGA2* mRNA was also detected by RT-PCR but not by northern blot analysis in three human fetuses of 19 to 22 weeks estimated gestational age by Gattas *et al*. [28]. Genbacev *et al*. [29] identified the chorionic mesoderm as a niche for human trophoblastic progenitor cells that support placental growth. *HMGA2* was found to be one of the factors associated with the self-renewal or differentiation of these cells.

The aim of the present study was to investigate temporal and spatial expression patterns of *HMGA2* in the human placenta aimed at insights into the role of HMGA2 in the development of this organ and into a possible use of this molecule as a biomarker indicating disturbances of proper implantation and placenta development.

## Methods

### Tissue specimens and RNA isolation

Formalin-fixed paraffin-embedded (FFPE) tissue samples were collected at the Institute for Pathology, Elbe Clinic Stade-Buxtehude, Germany. Pathological examinations were performed after haematoxylin and eosin staining of the samples. Six to eight sections of 5 μm for each sample were used for isolation of total RNA. Isolations were performed using the innuPREP Micro RNA Kit (Analytik Jena, Jena, Germany) according to the manufacturer’s instructions with the following modifications: Lysis of the paraffin sections preceding RNA isolation was conducted using TLS-Lysis Solution and Proteinase K from the innuPREP DNA Micro Kit (Analytik Jena) without prior deparaffinization. Sections were incubated for 1 h at 60°C and 15 min at 80°C.

### cDNA-synthesis and quantitative real-time RT-PCR

RNAs were reverse-transcribed into cDNA by M-MLV Reverse Transcriptase (Invitrogen, Karlsruhe, Germany). Quantitative real-time PCR (qRT-PCR) was performed using the Applied Biosystems 7300 sequence detection system according to Taq-Man Gene Expression Assay Protocol (Applied Biosystems, Darmstadt, Germany) in 96-well microtiter plates with a total volume of 20 μl. In the case of the TaqMan gene expression assay for *HMGA2* (assay number Hs00171569, Applied Biosystems, Foster City, USA), each reaction consisted of 2 μl of cDNA reverse transcribed from 25 ng of total RNA, 10 μl of TaqMan Universal PCR Master Mix (Applied Biosystems), 1 μl of TaqMan assay, and 7 μl of ddH_2_O. For the *HPRT* assay, using HPRT FP and HPRT RP primers [30], each reaction consisted of 2 μl of cDNA reverse transcribed from 25 ng of total RNA, 10 μl of TaqMan Universal PCR Master Mix, 600 nM (1.2 μl) of forward and reverse primers, and 200 nM (0.2 μl) of probe [30] and 5.4 μl of ddH_2_O. Thermal cycling conditions were 2 min at 50°C followed by 10 min at 95°C, 50 cycles at 95°C for 15 s and 60°C for 1 min. For each sample a negative control of the previous cDNA synthesis (missing reverse transcriptase) and for each plate a non-template control of amplification and a non-template control of previous cDNA synthesis were included in each run. Sequence Detection Software 1.2.3 (Applied Biosystems) was used to analyze data. All testing reactions were performed in triplicate. *HPRT* was used as endogenous control as it has previously been shown to be stably expressed in human fetal placenta [31-33]. As recommended for FFPE samples [34], the fragment sizes amplified by both assays were small, ranging between 65 and 80 bp. A validation of these values was performed via gel electrophoresis of the PCR product (data not shown). When applying the comparative C_T_ method, the sample with the lowest value was used as calibrator.

### Immunohistochemical analysis

Slides utilized for the immunohistochemical analysis were produced using consecutive sections directly adjacent to those for the qRT-PCR investigation. Immunohistochemical staining for HMGA2 (rabbit polyclonal anti-HMGA2-P3, Biocheck, Forster City, USA) was performed using a detection kit (DAKO, Glostrup, Denmark) and a semi-automated stainer (DAKO TechMate) according to the specifications of the manufacturer. For antigen retrieval the slides were treated in a PT Link module (DAKO) using the EnVision™ FLEX Target Retrieval Solution, low pH (DAKO). The antibody dilution used was 1:500.

A FFPE sample from a uterine leiomyoma carrying a *HMGA2* rearrangement leading to its overexpression served as positive control whereas negative control was performed by omission of the primary antibody. Staining extent was classified as negative, very weak, weak, moderate, and strongly positive, respectively.

### Statistical analysis

The two-sided Wilcoxon rank sum test to compare averages from two independent groups was used. Relationships between two observed or measured amounts were quantified by linear regression or by a nonparametric spline model. If the latter turned out to have a significantly better fit, models were compared by the likelihood ratio test. A *p-*value of less than 0.05 was considered significant, a *p-*value below 0.001 was deemed highly significant. Statistical calculations were done using the R package, version 2.3.2 [35].

### Ethics Statement

All samples investigated were initially taken for diagnostic purposes and only secondarily used for the present study. Samples were de-identified before their use in this study, in line with the rules of the Helsinki declaration. The study was approved by the local ethics committee (Aerztekammer Bremen: reference number 371).

## Results

### qRT-PCR analysis for the expression of *HMGA2* mRNA

Eighty-nine samples of human fetal placenta were tested for the expression of *HMGA2* mRNA (table 1A and 1B). Eighty-five were collected after termination of the pregnancy or abortion, respectively. In three cases no information was available about the type of abortion. In addition, four specimens from term placentae were examined as well. The placenta samples showed a relative expression ranging from 1 to 498 (fig. 1).

**Table 1:**
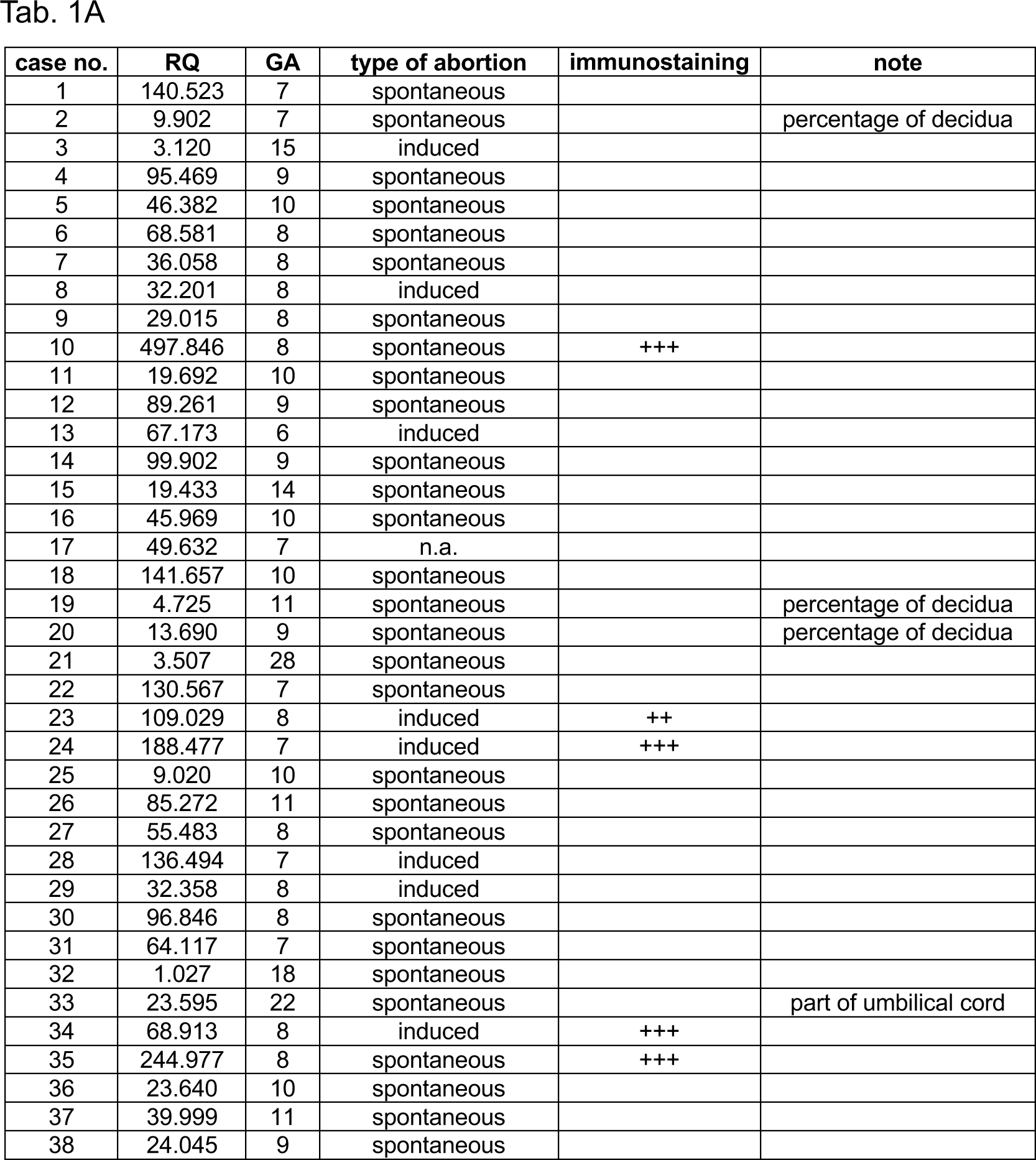
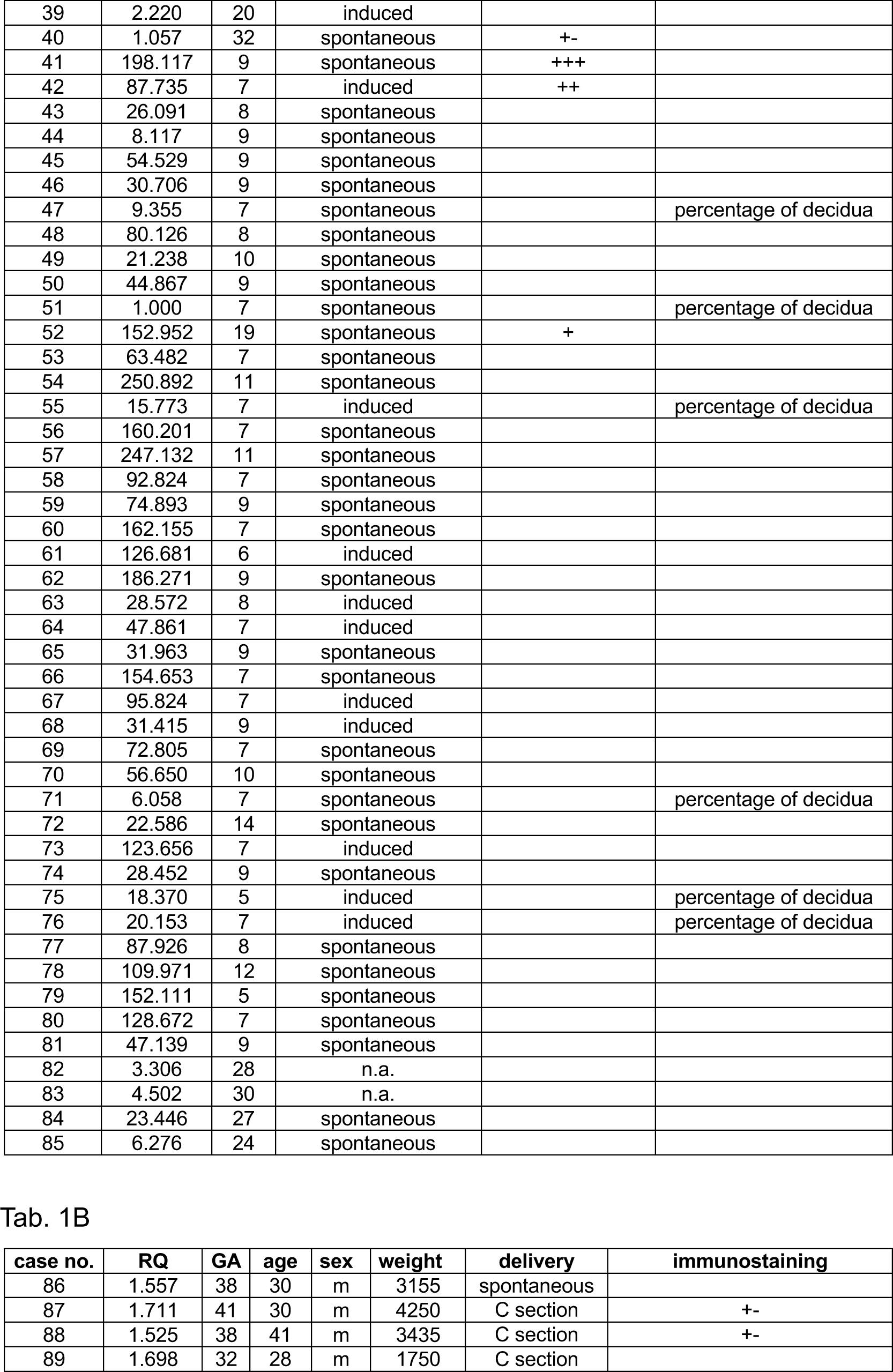
Summary of all samples investigated for the expression of *HMGA2*. RQ: relative quantification; GA: calendar gestational age in weeks; n.a.: information not available; percentage of decidua: sample consisted of at least 50 % decidua; umbilical cord: sample contained a section of the umbilical cord; immunostaining: sample was used for immunostaining with an HMGA2-specific antibody. Staining extent: +-very weak. + weak. ++ moderate. +++ strong age: age of the mother at delivery; sex: sex of the neonate; weight: weight of the neonate in grams; C section: Caesarean section A: samples collected after abortion or stillbirth B: samples collected after birth

**Figure 1:**
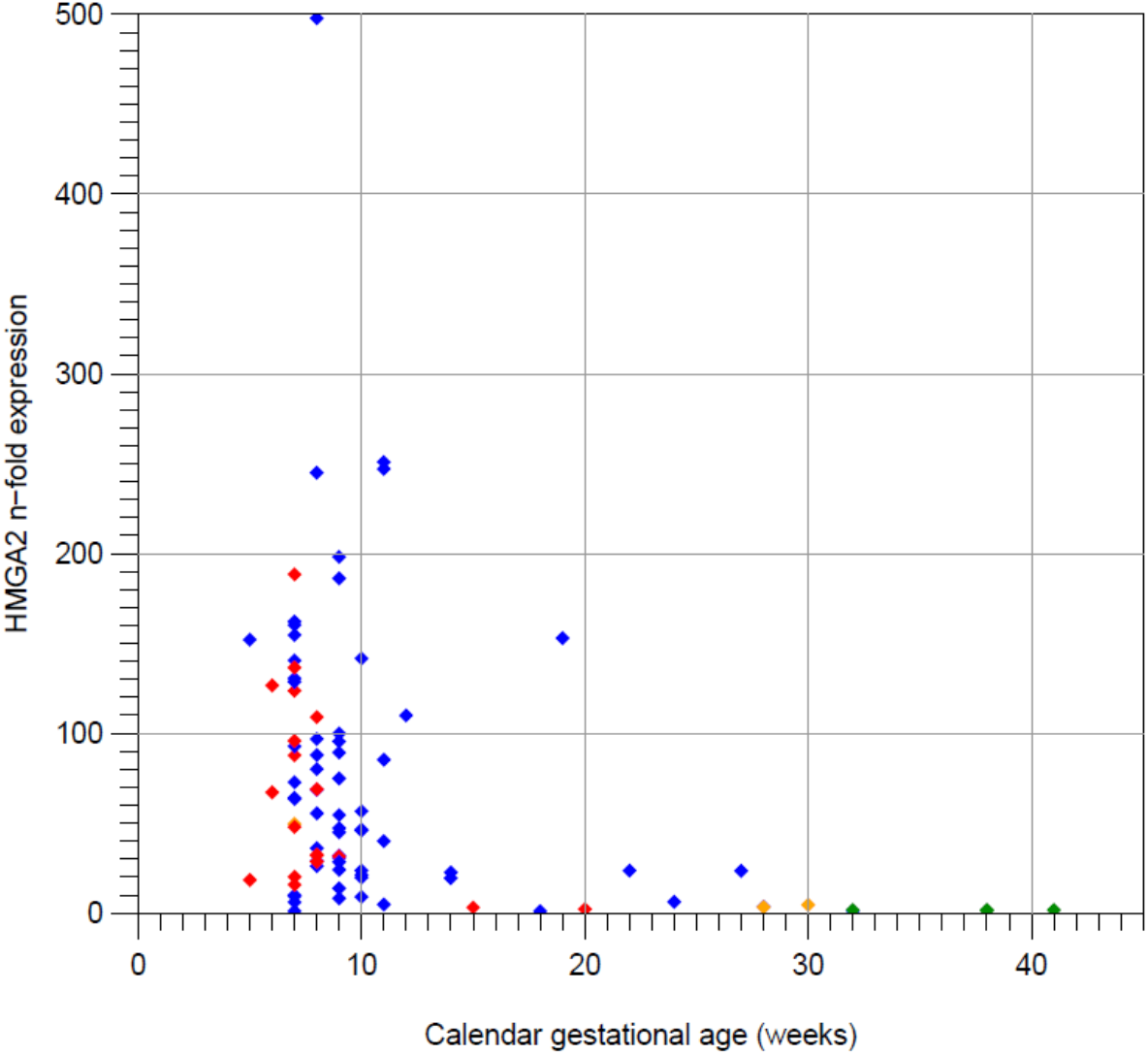
*HMGA2* expression in relation to the gestational age. Linear display for *HMGA2* expression, all samples. Blue: spontaneous abortion, red: induced abortion, green: gathered postpartum, orange: abortion, no information available on type of abortion.

Overall, a strong decline of the expression of *HMGA2* was noted with gestational age. For all placenta samples, the correlation coefficient was 0.653 (*p* = 2.37*10^−12^) (fig. 2). When the analysis was restricted to the specimens after induced abortion (IA) and after delivery (AD), the *r-*value was 0.889 (*p* = 3.88*10^−9^). The samples collected after spontaneous abortion (SA) showed a correlation coefficient of 0.431 (*p* = 1.30*10^−4^). The relation between gestational age and *HMGA2* expression did not significantly differ between these groups (*p* = 0.095, likelihood ratio test).

**Figure 2:**
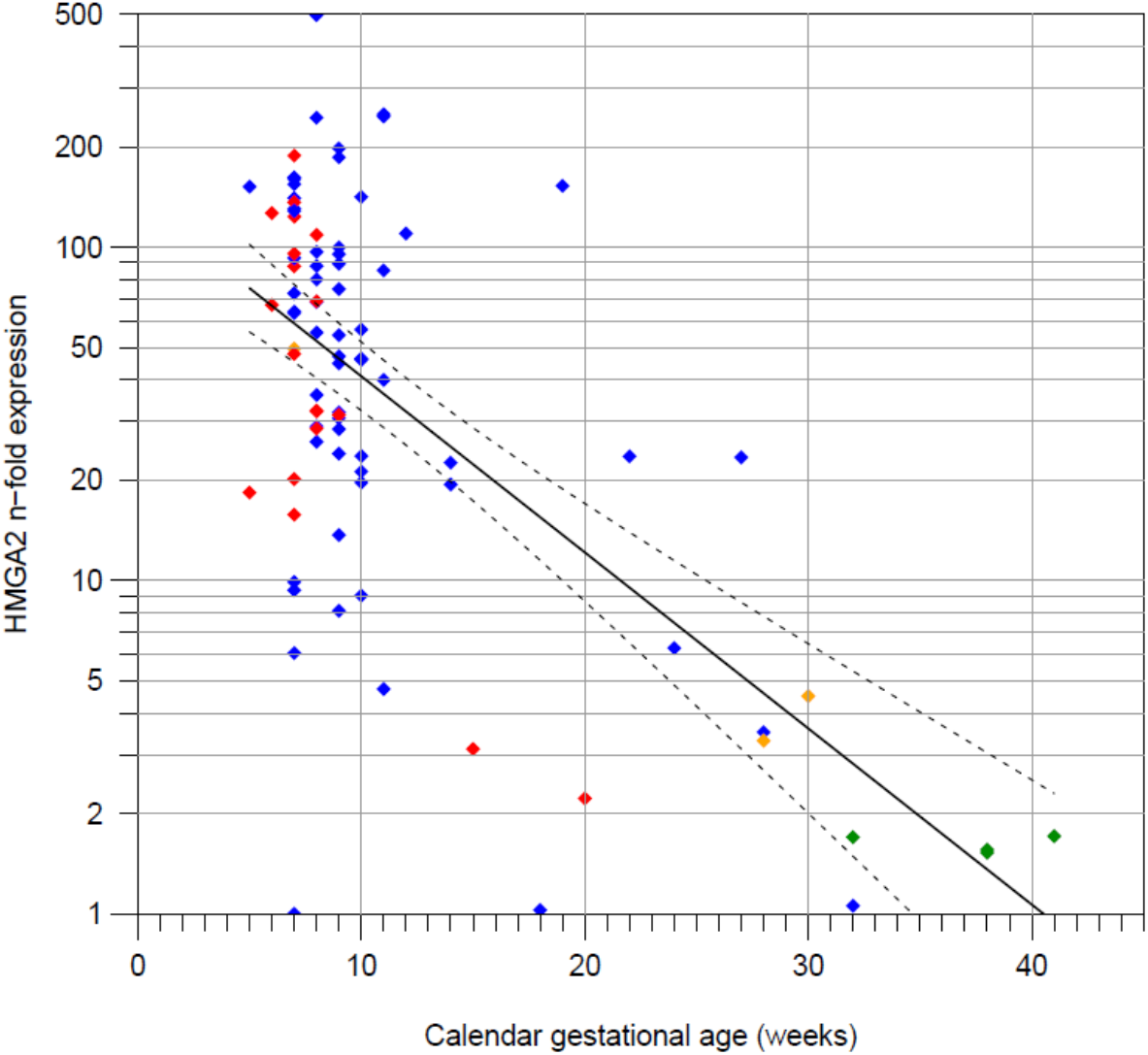
*HMGA2* expression in relation to the gestational age including the linear regression line with 95% confidence range. Logarithmic display for *HMGA2* expression, all samples. Coloring of the rhombi denotes the type of sample. Blue: spontaneous abortion, red: induced abortion, green: gathered after delivery, orange: abortion, no information available on the type of abortion.

*HMGA2* mRNA levels from samples taken during the first trimester of pregnancy were significantly higher than those of the second and third trimester (p = 3.82*10^−7^, Wilcoxon signed rank test). Using the same test, significant differences were also found within the SA subgroup (p = 0.00257) and within the IA and AD joint subgroups (p = 1.98 * 10^−5^). Overall, a relatively wide ranging but overall high level of expression was observed up to 13 weeks (calendar gestational age (GA)). After that, the level of expression strongly declined up to a CGA of 28 weeks where it finally reached a steady value for the rest of the pregnancy. In specimens of induced abortions, the observed drop was more pronounced and happened earlier, at around nine weeks of gestation.

Pathological examinations of the specimens after haematoxylin and eosin staining revealed a considerable percentage of maternal decidua in some of the samples (see also table 1). After immunohistochemical analysis (see also below), the decidua was found to be HMGA2 negative (data not shown). Nevertheless, omitting this data does not change the general expression patterns of *HMGA2* during pregnancy.

### Immunohistological analysis

Eleven samples were investigated for the presence and localization of HMGA2 by immunohistochemical analysis. In general, samples from early gestation (eight to ten weeks GA) showed intense signals (fig. 3A) with the strongest staining visible in the nuclei of the stroma cells and slightly less intense signals in the cytoplasm of the trophoblast. In contrast, samples from GA between 19 and 41 weeks showed only weak to very weak signals (fig. 3B). Overall, the results from the immunostaining are in concordance with those from the qRT-PCR, except for two outliers. Decidual tissue as found in some of the samples turned out to lack detectable expression of HMGA2 by immunostaining.

**Figure 3:**
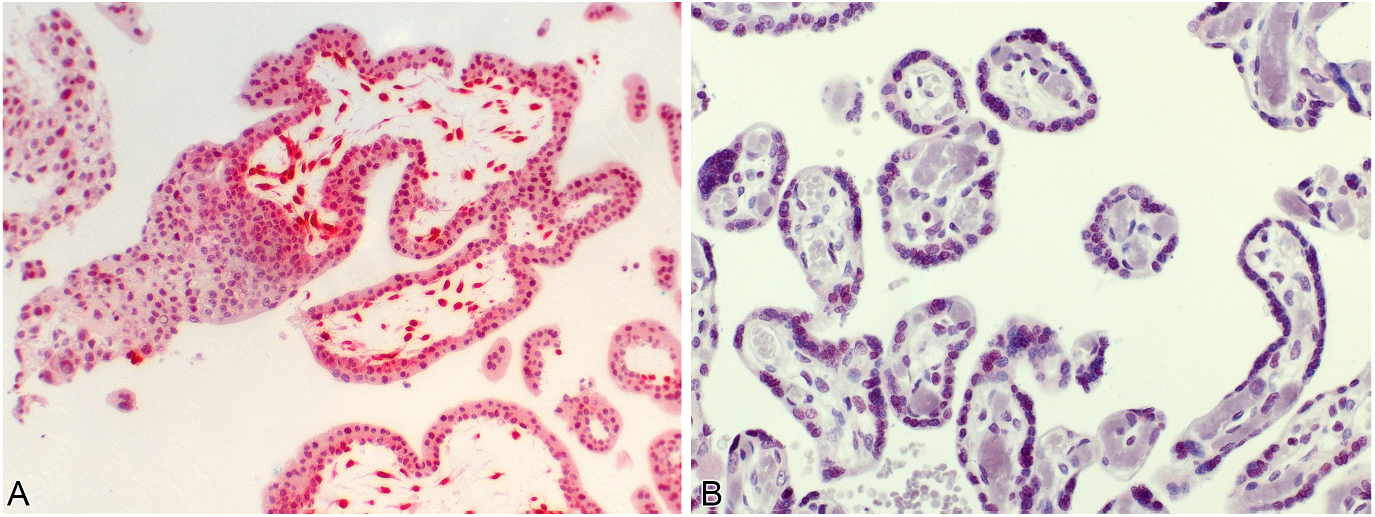
Immunoreactivity for HMGA2. In all but two cases the qRT-PCR data was in concordance with the interpretation of the HMGA staining. (A) Seven weeks calendar gestational age showing an intensive signal (red staining) for HMGA2 and a high expression as measured in the qRT-PCR. (B) Case no. 88 (38 weeks calendar gestational age) with a barely visible signal, the qRT-PCR showed a very low expression. In all samples that stained positive, HMGA2 was found in the nuclei of the stroma cells of the villi and, with a much weaker intensity, in the cytoplasm of the trophoblast.

## Discussion

A highly significant decrease of the level of *HMGA2* mRNA with increasing CGA was found in this study. The level of *HMGA2* declines towards the end of the first trimester with an apparently stable but low level reached around week 28. In samples from induced abortions, the *HMGA2* level dropped around the ninth week of CGA. In induced and spontaneous abortions as one group, the decrease is less steep and takes place several weeks later. For the specimens collected after spontaneous abortion it is conceivable that the underlying cause of the abortion had affected the development of the placenta and the expression of *HMGA2*. In addition, the embryo or fetus might have died a varying period of time before abiosis of the placenta [36,37]. Therefore, despite a smaller sample size, more reliable results may be obtained from the group of specimens gathered after induced abortion or after birth. The stable level of *HMGA2*-expression detected in samples from the third trimester as well differs from the analysis by Rogalla *et al*. [8] revealing an absence of *HMGA2*-expression in term placentae. A possible explanation is the higher sensitivity of qRT-PCR compared to conventional RT-PCR.

*HMGA2* is known to participate in the proliferation of tissues by upregulation of genes involved in cell proliferation and invasion [reviewed in [1,38]. Therefore, the results presented in this study are in accordance with those published by Sitras *et al*. [39] who tested more than 29.000 genes in human placentas. Applying microarray analysis, they found that genes involved in cell proliferation, differentiation, and angiogenesis are upregulated in human first trimester placentae. In this latter study, the expression of HMGA2 was not tested.

As demonstrated here, high *HMGA2* expression correlates with the uterine low oxygen environment in early pregnancy. As a result of trophoblast invasion into the maternal decidua, spiral arteries are plugged during the first 7 to 8 weeks of pregnancy [reviewed in [40]]. Filtrated plasma enriched with secretions from the endometrial glands can be found in the placental intervillous space, providing histiotrophic nutrition [41]. A low oxygen environment has been shown to be necessary for the proliferation of cytotrophoblast cells [42,43]. Around week 7 to 8 of gestation (week 9 to 10 CGA), maternal uterine circulation to the placenta begins [44], resulting from disintegration of spiral arterial plugs [45], indicating the starting of haemotrophic nutrition of the fetus, which is reaching its full function from week 12 of gestation on. The increasing oxygen level coincides with the decrease of *HMGA2* expression.

Spatially and temporally accurate proliferation and invasion of trophoblast cells are crucial for an undisturbed pregnancy. Superficial implantation of the placenta leading to poor placental and uterine perfusion is characteristic for preeclampsia [46], reviewed in [47]. Preeclampsia affects 2 % to 8 % of all pregnancies [48] and is the cause of direct maternal death in 16 % [49] to 39 % of cases [50]. The only treatment of this serious medical condition is planned preterm delivery. Preeclampsia is not yet fully understood and there are few markers for diagnosis [51]. In combination with oxidative stress of the placenta [reviewed in [36]], several transcription factors involved in the proliferation and differentiation of the trophoblast have been detected to contribute to an elevated risk of preeclampsia [52-55]. For some proteins a significantly higher expression has been shown in early gestation [53,54,56], similar to the results for *HMGA2*. While symptoms of preeclampsia do not appear before the 20th week of gestation, it seems to result from earlier changes of proliferation and differentiation of the trophoblast that play a key role in the implantation of the embryo during low placental oxygenation [reviewed in [57]. In this study, in case number 89 a severe form of preeclampsia leading to a premature delivery at the gestational age of 32 weeks (see also table 1) was diagnosed. The measured value for *HMGA2* was within the normal range of probes of this late stage of pregnancy. In case of a correlation between *HMGA2* and preeclampsia, a deviation might be restricted to the first trimester since in that period of time the proliferation and invasion of the trophoblast determines whether an elevated risk for preeclampsia will exist. A similar situation has been proposed by Jeon *et al*. [51] for IMUP-2. They suggest an association of this protein with preeclampsia but in term placentae their findings do not reveal differences for patients with or without preeclampsia.

Whereas shallow infiltration of the trophoblast is a sign for preeclampsia, overly deep infiltration indicates another obstetric complication: placenta accreta (including the closely related forms of increta and percreta). This severe complication during pregnancy has been associated with decidual deficiency and an overinvasive trophoblast (reviewed in [58]). Placenta accreta accounts for about 1 % of maternal mortality in the USA [36] and approximately 5 % of the women with this complication die [59-61]. In addition, fetal deaths occur in almost 26 % of the cases [61]. Since the 1970s the incidence has risen from one in approximately 4,000 deliveries to one in 333 [59,60,62,63]. The reasons are unknown, making further investigations necessary. Since there is no definitive method [64] to detect this complication before birth, a possible test would be of high interest. However, aberrant *HMGA2* expression cannot be determined by a screening procedure using non-invasive methods. Further investigations are needed to check if determination of HMGA2 or its mRNA in maternal serum might be a reliable method.

By immunostaining it became apparent that HMGA2 is strongly expressed in the stroma cells of the placental villi. Positive staining mainly was noted in the nucleus, but in a lesser concentration also observed in the cytoplasm of the trophoblast. This pattern is significantly different from that of HMGA1 [65] found exclusively in the trophoblast cells. This suggests different roles for the two members of the HMGA family. The results of this study are in concordance with those from Genbacev *et al*. [66] who also found HMGA2 in the villi as well as a change from nuclear to cytoplasmatic location. In general, HMGA proteins are considered nuclear proteins [1], even though cytoplasmatic expression of HMGA1 has been reported before [65]. Thus, in the majority of papers only nuclear immunoreactivity for HMGA2 was noted [67,68].

## Data Availability

available upon request

## Authors’ contributions

Study design: LB, JB

Provision of sample, immunostaining for HMGA2, clinical workout: BMH

Molecular genetic studies: LB

Pathological determination of samples: BMH, AG Immunohistochemical analysis: BMH, LB, AG

Statistical analysis and interpretation of data: WW, LB, WK, JB

Writing of manuscript: LB, AG, WK, JB

All authors have read and approved the final manuscript.

## Relevant ethical guidelines

All relevant ethical guidelines have been followed, all necessary IRB and/or ethics committee approvals have been obtained, all necessary patient/participant consent has been obtained and the appropriate institutional forms archived.

